# Phenome-Wide Risk Evaluation of GLP-1 Receptor Agonist Use in Type 2 Diabetes with Real-World Data Across Multiple Healthcare Systems

**DOI:** 10.1101/2025.08.13.25333579

**Authors:** Rohit Vashisht, Sanket S. Dhruva, Ayan Patel, Lisa Dahm, Pagan Morris, David Gonzalez, Rob Follett, Marina Sirota, Karandeep Singh, Cora Han, Suneil Koliwad, Atul J. Butte

**Author notes:** **Correspondence:** Rohit Vashisht, PhD, Bakar Computational Health Sciences Institute University of California, San Francisco, UCSF Valley Tower, Box 2993 490 Illinois Street, Floor 2 San Francisco, CA, 94143.

## Abstract

Glucagon-like peptide-1 receptor agonists (GLP-1RAs), widely used for managing type 2 diabetes (T2D) and weight, are gaining attention for treatment of a broad set of conditions. Large-scale, real-world evidence of their broader clinical impact is needed. We assessed the phenome-wide risks and benefits of new users of GLP-1RA versus sulfonylureas, SGLT-2 inhibitors (SGLT-2i), and DPP-4 inhibitors (DPP-4i) in adults with T2D in a multi- healthcare center, 1:1 high-dimensional propensity score matched new user cohort study utilizing principles of target trial emulation with electronic health records (EHRs) of over 9 million patients followed up to 730 days. Primary outcomes included 239 clinical endpoints across 14 organ systems. Among 86,790 patients, mean age was 58-66 years and 44-62% were female across cohorts. Compared to DPP-4i, GLP-1RA use was associated with a reduced risk of acute myocardial infarction (sHR 0.61, 95% CI 0.43–0.85) and chronic kidney disease (sHR 0.71, 95% CI 0.62–0.81). Compared to sulfonylurea, GLP-1RA use was associated with reduced acute renal failure risk (sHR 0.77, 95% CI 0.65–0.90). GLP-1RA use was associated with reduced heart failure risk compared to SGLT-2i (sHR 0.66, 95% CI 0.55–0.80). GLP-1RA also was associated with lower epilepsy risk versus DPP-4i (sHR 0.49, 95% CI 0.32–0.76). GLP1RA was associated with elevated risks of nausea/vomiting: sHR 1.37 vs SGLT2i, 95% CI 1.15–1.62; sHR 1.46 vs sulfonylureas, 95% CI 1.24–1.71). Head-to-head real-world comparisons with established T2D therapies confirmed the broad cardiorenal and metabolic benefits and known on-target adverse effects of GLP-1RAs –consistent with randomized clinical trials - but also suggest potential risks for musculoskeletal and genitourinary adverse events, warranting continued real-world post-market surveillance.

## Introduction

Glucagon-like peptide-1 receptor agonist (GLP-1RA) use among U.S. adults with type 2 diabetes (T2D) surged to 19.4% by 2022, representing 5.1 million people and quadrupling from 2016 levels^1,2^. While gastrointestinal adverse effects of GLP-1RAs are well-established^3^, their adoption has extended beyond glycemic control to include indications such as weight management^1^, driven by accumulating evidence of pleiotropic effects. These benefits include reductions in cardiovascular mortality, renal disease progression, systemic inflammation, hepatic fibrosis, and hospitalization rates^4–17^. Additionally, emerging data suggest a potential protective association with Alzheimer disease^18^.

As real-world evidence accumulates, it is increasingly clear that GLP-1RA confer both substantial clinical benefits and potential risks on a multitude of clinical endpoints^19^. Therefore, a rigorous and comprehensive appraisal of GLP-1RA utilization across heterogeneous populations and its correlation with a broad spectrum of clinical endpoints in multiple healthcare systems would provide evidence about safety and effectiveness of this medication class. Such systematic investigations are pivotal for refining clinical practice guidelines, optimizing therapeutic strategies, guiding subsequent mechanistic and interventional research, and ultimately enabling evidence-based clinical decision-making.

Accordingly, building on our earlier framework^20^, we utilized the principles of target trial emulation framework to systematically and comprehensively characterize the phenome-wide safety and effectiveness profiles of GLP-1RA relative to sulfonylureas, sodium-glucose cotransporter-2 inhibitors (SGLT-2i), and dipeptidyl peptidase-4 inhibitors (DPP-4i) among individuals with T2D across five large, diverse U.S. healthcare systems in the state of California^20,21^.

## Methods

### Ethical Statement

The Institutional Review Boards across the UC Health system determined that the research use of the HIPAA limited data set for this cohort study did not constitute human participants research and was therefore exempt from further approval and informed consent. This study is reported in accordance with the Strengthening the Reporting of Observational Studies in Epidemiology (STROBE) guidelines. The analysis was conducted between October 2024 and April 2025 using electronic health record data beginning in 2012.

### Data Source and Study Design

Data for this study were derived from the University of California Health Data Warehouse (UCHDW), a Health Insurance Portability and Accountability Act (HIPAA)-compliant, de-identified electronic health record (EHR) repository encompassing over 9 million patients across a longitudinal span of 13 years. The UCHDW integrates data from six independent academic medical centers—UC San Francisco, UC San Diego, UC Davis, UC Irvine, UC Los Angeles, and UC Riverside—representing 12 affiliated hospitals with approximately 150,000 inpatient and 4 million outpatient visits annually. Given its relatively recent establishment and limited cohort of patients with type 2 diabetes (T2D), UC Riverside was excluded from the analysis. The remaining five sites were anonymized as UC-1 through UC-5 for analytical purposes. All data were harmonized using the Observational Medical Outcomes Partnership Common Data Model (OMOP-CDM), facilitating standardized extraction, transformation, and loading (ETL) procedures across institutions^20,22^. The analytic cohort included patients initiating glucose-lowering therapies between January 2012 and December 2024, with analyses conducted from October 2024 to April 2025. Patients were followed up to 730-day period post-index treatment initiation to assess longitudinal outcomes.

An overview of the study design is illustrated in **Figure-1**. Building on our previous work^20^, we implemented a structured, multi-phase analytical pipeline **(Figure-1)**. First, within each healthcare system, we identified new-user cohorts of individuals initiating GLP-1RA, sulfonylurea, SGLT-2i or DPP-4i, **(Figure-1 A-C)**. Second, we applied high-dimensional propensity score matching to construct pairwise, site-specific treatment and comparisons cohorts that were balanced on a broad array of baseline covariates **(Figure-1 D-E)**. Third, we conducted a systematic, longitudinal assessment of associations between GLP-1RA use and over 200 clinical endpoints at human phenome-wide scale spanning across 14 organ systems, using up to a 730-day follow-up period after treatment initiation **(Figure-1 F)**. Fourth, we synthesized site-specific estimates via meta-analysis for each drug-pair comparison and evaluated the robustness of the pooled estimates using a leave-one-medical-center-out (LOMCO) sensitivity analysis **(Figure-1 G-H)**. Finally, we performed quantitative bias analyses to assess the potential influence of unmeasured confounding and applied multiple comparison adjustments to yield a statistically rigorous, hierarchically structured map of real-world treatment associations **(Figure-1 I-J)**.

**Figure 1:**
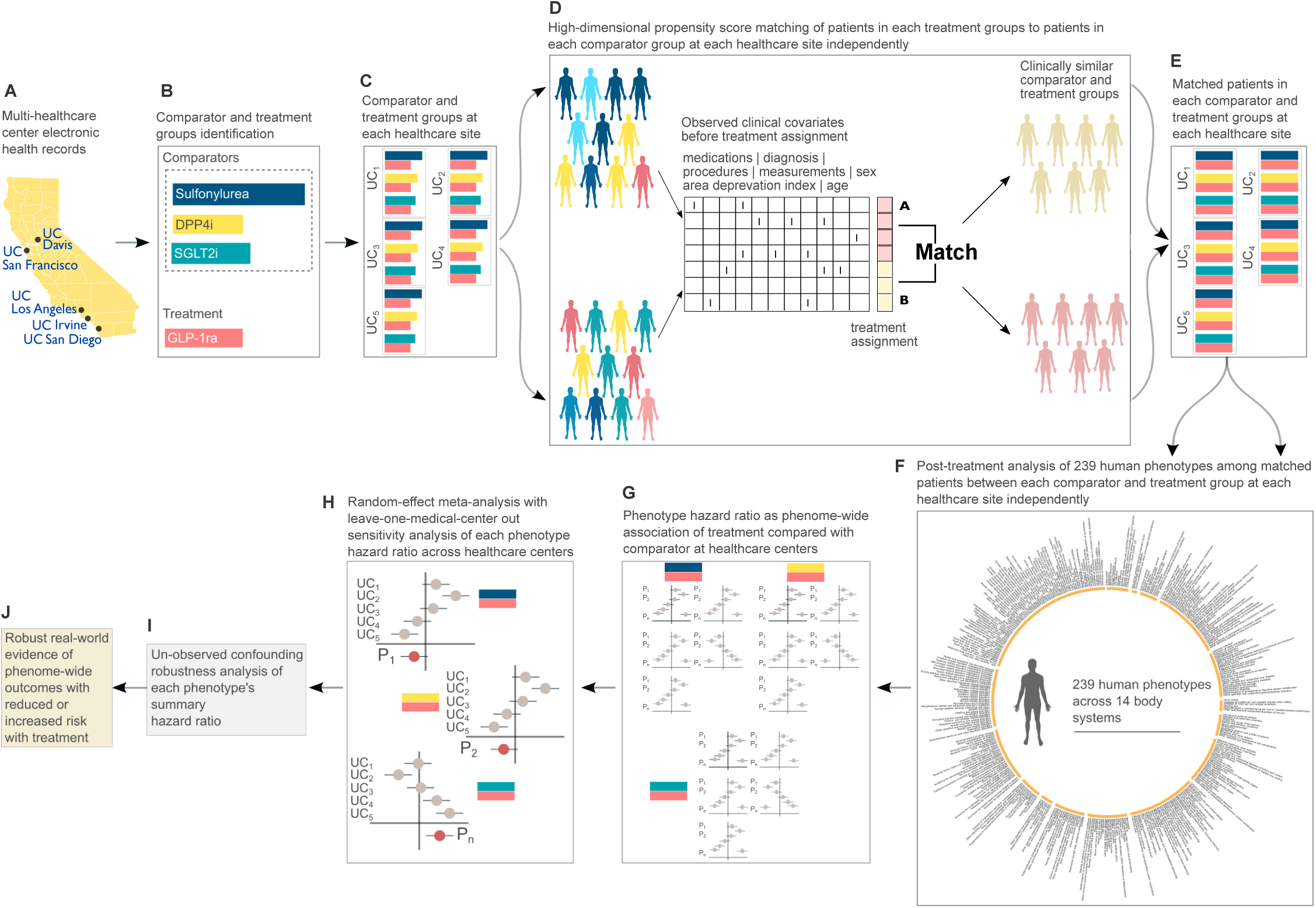
Overview of the study design. (A) Real-World data source across UC Health B) Treatments considered in this study c) Cohort identification following inclusion and exclusion criteria (D) High-dimension propensity score estimation of treatment assignment at each site individually (E) Matched cohorts of each comparator and treatment at each site (F-G) Phenome-wide outcome analysis of matched cohort at each site independently (H) Random-effect meta-analysis with leave-one-medical-center-out stability analysis of each effect estimate across UC Health (I) Analysis of the robustness of the summary estimates with respect to un-measured confounding (J) Evidence landscape at varying degree of statistical certainty, significance and un-measured confounding.

### New User Cohorts

New-user cohorts of GLP-1RA, sulfonylurea, SGLT-2i and DPP-4i were defined based on standardized inclusion and exclusion criteria. For each drug class, initiation was determined by the first recorded prescription order in the EHR, which served as the index date. Taking the GLP-1RA cohort as an illustrative example, patients were excluded if they had prior prescription for a comparator agent (sulfonylurea, SGLT-2i or DPP-4i) on or any time before the index date, ensuring treatment naivety. Additional exclusion criteria were: (1) any recorded diagnosis of type 1 diabetes or secondary diabetes (including gestational diabetes) at or any time prior to the index date; (2) absence of a diagnosis of type 2 diabetes prior to or within 7 days following the index date; (3) age < 18 years on the index date; and (4) lack of continuous enrollment for a minimum of 30 days preceding treatment initiation, defined as not having uninterrupted health insurance coverage during the 30-day period leading up to the start of treatment. This criterion was applied to confirm that individuals had consistent access to healthcare services prior to initiating therapy within UC Health. A detailed accounting of cohort construction and attrition, including the number of patients satisfying inclusion and exclusion criteria for each medication group at each UC site, is provided in **eTable-1**.

### Propensity Score Estimation, Matching and Covariate Balance Diagnostics

Propensity scores (PS), defined as the conditional probability of receiving the treatment of interest conditional on observed covariates, were estimated separately for each treatment cohort (Tc)–comparator cohort (Cc) pair using L1-penalized logistic regression via the least absolute shrinkage and selection operator (LASSO). Ten-fold cross-validation was employed to identify the optimal regularization parameter, with λ.1se (the largest λ within one standard error of the minimum cross-validation error) selected to maximize model sparsity while preserving predictive performance^23^. To construct the high-dimensional covariate space, a binary patient feature design matrix (PFDM) was generated for each Tc–Cc pair. Rows represented individual patients in the treatment and comparator arms, and columns encoded covariates captured within the 365-days prior to the index date, including: age at index date, sex, comorbidities, prior diabetes medication use (including insulin and metformin), laboratory testing, procedural history, number of healthcare encounters (as a proxy for healthcare utilization), and area deprivation index (as a proxy for neighborhood-level socioeconomic status). Following PS estimation, 1:1 nearest-neighbor matching without replacement was performed using a caliper of 0.20 standard deviations on the logit-transformed PS. Balance diagnostics were evaluated using absolute standardized mean differences (SMDs), with covariates achieving |SMD| < 0.10 post-matching considered adequately balanced^24^. Matching procedures were executed independently for each Tc–Cc comparison within each UC Health site to preserve site-level heterogeneity. All analyses were implemented in R version 3.5.0 within the Databricks analytics environment version 16.3^25^.

### Outcomes

Outcome definitions were obtained from the Clinical Classifications Software Refined (CCSR) version 2025.1 (https://hcup-us.ahrq.gov/toolssoftware/ccsr/ccs_refined.jsp), developed as part of the Healthcare Cost and Utilization Project (HCUP)—a federal-state-industry initiative sponsored by the Agency for Healthcare Research and Quality (AHRQ). The outcomes spanning 14 body systems: endocrine and metabolic, nervous system, genitourinary system, neoplasms, respiratory system, blood and blood-forming organs, skin and subcutaneous tissue, eye and adnexa, circulatory system, musculoskeletal and connective tissue, symptoms, digestive system, mental health, and ear and mastoid process were considered. To comply with HIPAA requirements for de-identified data across UC Health, outcomes with fewer than 10 events during follow-up in any study arm were excluded. The final set comprised 239 outcomes across 14 body systems.

### Statistical Analysis

Cox proportional hazards regression was used to estimate hazard ratios (HR) quantifying the comparative risk of outcomes associated with initiation of GLP-1RA versus each active comparator (sulfonylurea, SGLT-2i, DPP-4i) within propensity score–matched Tc-Cc pairs at each UC Health site independently. Follow-up commenced on day 1 post-treatment initiation to mitigate immortal time bias^26^ as well as day 0 inflation bias^27^ associated with index-day events and continued for 730 days unless censored. Patients were censored at the earliest occurrence of treatment discontinuation to an agent outside the designated Tc–Cc pair or end of the observation window. Individuals with a history of the outcome of interest prior to treatment initiation were excluded to ensure incident outcome analysis.

Site-specific HRs, along with corresponding 95% confidence intervals (CIs) and p-values, were estimated using the Cox model. To synthesize evidence across sites, a random-effects meta-analysis was conducted for each Tc–Cc comparison using a restricted maximum likelihood (REML) estimator, yielding summary hazard ratios (sHR). Between-site heterogeneity was assessed using the I² statistic, representing the proportion of variance attributable to heterogeneity rather than sampling error^28^. The stability of sHR was further evaluated via leave-one-medical-center-out (LOMCO) influence analysis. A summary estimate was deemed stable if the point estimate remained within the 95% CI of the sHR following exclusion of any single UC Health site. The robustness of each summary hazard ratio (sHR) to potential unmeasured confounding was further evaluated by calculating E-values, which quantify the minimum strength of association that an unmeasured confounder would need to have with both treatment and outcome to fully explain the observed association^29^.

To systematically classify the strength of evidence, we implemented a hierarchical inferential framework – a structured, multi-stage approach that progressively applies increasingly stringent criteria in sequence to support hypothesis generation and provide evidence that could inform clinical decision-making. Associations were considered hypothesis-generating if the 95% CI for the sHR excluded the null (HR = 1). For findings to be considered stable, robust and potentially clinically actionable, three criteria were required: (1) sHR stability under LOMCO analysis, (2) E-value ≥ 1.5, and (3) false discovery rate–adjusted p-value < 10%.

## Results

### Population Characteristics

Across the UC health system, sulfonylureas were the most frequently initiated therapy among analyzed patients, with 28,012 new users, followed by GLP-1RA (22,853 users), SGLT-2i (19,242 users), and DPP-4i (16,683 users) **(Table 1)**. New users of GLP-1RA were younger on average (mean age 56.9–59.1 years) compared with those initiating SGLT-2i (63.8–66.4 years), DPP-4i (63.8–65.9 years), or sulfonylurea (60.5–62.7 years). There were differences by sex in medication: new GLP-1RA users were predominately female (56%–62%), whereas SGLT-2i were more commonly initiated in male patients (59%–62%). Baseline healthcare utilization varied by medication class. SGLT-2i users had the highest utilization (62.8–109.9 visits), followed closely by GLP-1RA users (66.4–96.8 visits). In contrast, sulfonylurea users demonstrated the lowest utilization (21.0–32.9 visits).

**Table 1:** Population Characteristics. Baseline clinical conditions were identified based on standardized definitions of clinical phenotypes corresponding to components of the Charlson Comorbidity Index. Socioeconomic status was assessed using the Area Deprivation Index (ADI), categorized into deciles (10-percentile intervals) according to established methodology^46^. Patient reported sex was used, and age was calculated on the index date. Baseline healthcare utilization included any type of clinical visit.

| Characteristics |  | UC-1 | UC-2 | UC-3 | UC-4 | UC-5 |
| --- | --- | --- | --- | --- | --- | --- |
| **Dipeptidyl peptidase-4 inhibitors** |  |  |  |  |  |  |
| New users, No. |  | 2820 | 2305 | 4741 | 3194 | 3623 |
| Sex, No. | Female | 1356 (48.09%) | 1063 (46.12%) | 2217 (46.76%) | 1566 (49.03%) | 1772 (48.91%) |
|  | Male | 1464 (51.91%) | 1242 (53.88%) | 2524 (53.24%) | 1628 (50.97%) | 1851 (51.09%) |
| Age, mean (SD) |  | 63.8 (12.8) | 65.3 (12.8) | 65.4 (13.1) | 65.9 (13.3) | 64.1 (12.6) |
| Treatment Discontinuation, NO. | Yes | 1289 | 917 | 2068 | 1241 | 1580 |
|  | No | 1531 | 1388 | 2673 | 1953 | 2043 |
| Baseline healthcare visits, mean (SD) |  | 39.8 (78.6) | 44.8 (77.9) | 52.2 (98.2) | 41.5 (78.2) | 47.8 (83.2) |
| Area deprivation index, No. % | 1 | 13 (0.5%) | 492 (21.3%) | 1111 (23.4%) | 188 (5.9%) | 197 (5.4%) |
|  | 2 | 46 (1.6%) | 512 (22.2%) | 616 (13%) | 261 (8.2%) | 209 (5.8%) |
|  | 3 | 113 (4%) | 205 (8.9%) | 684 (14.4%) | 479 (15%) | 253 (7%) |
|  | 4 | 165 (5.9%) | 168 (7.3%) | 494 (10.4%) | 400 (12.5%) | 483 (13.3%) |
|  | 5 | 178 (6.3%) | 106 (4.6%) | 509 (10.7%) | 557 (17.4%) | 412 (11.4%) |
|  | 6 | 250 (8.9%) | 127 (5.5%) | 409 (8.6%) | 540 (16.9%) | 408 (11.3%) |
|  | 7 | 377 (13.4%) | 107 (4.6%) | 303 (6.4%) | 304 (9.5%) | 504 (13.9%) |
|  | 8 | 498 (17.7%) | 157 (6.8%) | 177 (3.7%) | 168 (5.3%) | 365 (10.1%) |
|  | 9 | 664 (23.5%) | 163 (7.1%) | 124 (2.6%) | 106 (3.3%) | 216 (6%) |
|  | 10 | 403 (14.3%) | 188 (8.2%) | 142 (3%) | 120 (3.8%) | 401 (11.1%) |
|  | NA | 113 (4%) | 80 (3.5%) | 172 (3.6%) | 71 (2.2%) | 175 (4.8%) |
| Baseline comorbidities, No. % | Myocardial infarction | 196 (7%) | 158 (6.9%) | 237 (5%) | 193 (6%) | 297 (8.2%) |
|  | Congestive heart failure | 332 (11.8%) | 349 (15.1%) | 450 (9.5%) | 362 (11.3%) | 647 (17.9%) |
|  | Cerebrovascular disease | 238 (8.4%) | 234 (10.2%) | 321 (6.8%) | 295 (9.2%) | 276 (7.6%) |
|  | Dementia | 85 (3%) | 95 (4.1%) | 136 (2.9%) | 147 (4.6%) | 126 (3.5%) |
|  | Chronic pulmonary disease | 412 (14.6%) | 398 (17.3%) | 574 (12.1%) | 305 (9.5%) | 581 (16%) |
|  | Rheumatologic disease | 67 (2.4%) | 75 (3.3%) | 138 (2.9%) | 81 (2.5%) | 97 (2.7%) |
|  | Peptic ulcer disease | 78 (2.8%) | 67 (2.9%) | 142 (3%) | 70 (2.2%) | 158 (4.4%) |
|  | Mild liver disease | 214 (7.6%) | 272 (11.8%) | 437 (9.2%) | 214 (6.7%) | 560 (15.5%) |
|  | Hemoplegia or paraplegia | 62 (2.2%) | 61 (2.6%) | 51 (1.1%) | 71 (2.2%) | 51 (1.4%) |
|  | Renal disease | 789 (28%) | 846 (36.7%) | 1202 (25.4%) | 948 (29.7%) | 1368 (37.8%) |
|  | AIDS/HIV | 12 (0.4%) | 45 (2%) | 21 (0.4%) | 25 (0.8%) | 58 (1.6%) |
|  | Metastatic solid tumor | Less than 10 | Less than 10 | Less than 10 | Less than 10 | Less than 10 |
|  | Moderate to severe liver disease | 56 (2%) | 114 (4.9%) | 181 (3.8%) | 62 (1.9%) | 323 (8.9%) |
|  | Any malignancy | 423 (15%) | 527 (22.9%) | 905 (19.1%) | 552 (17.3%) | 689 (19%) |
| **GLP-1 receptor agonists** |  |  |  |  |  |  |
| New users, No. |  | 3669 | 3682 | 6802 | 2807 | 5893 |
| Sex, No. | Female | 2274 (61.98%) | 2125 (57.71%) | 3985 (58.59%) | 1706 (60.78%) | 3309 (56.15%) |
|  | Male | 1395 (38.02%) | 1557 (42.29%) | 2817 (41.41%) | 1101 (39.22%) | 2584 (43.85%) |
| Age, mean (SD) |  | 57.5 (12.9) | 58.1 (14.1) | 56.9 (13.7) | 57.1 (14.2) | 59.1 (12.9) |
| Treatment Discontinuation, NO. | Yes | 639 | 595 | 1072 | 596 | 1097 |
|  | No | 3030 | 3087 | 5730 | 2211 | 4796 |
| Baseline healthcare visits, mean (SD) |  | 96.8 (123.4) | 75.5 (114.5) | 92.7 (130.8) | 66.4 (103.2) | 92.2 (140.8) |
| Area deprivation index, No. % | 1 | 11 (0.3%) | 1047 (28.4%) | 1531 (22.5%) | 251 (8.9%) | 484 (8.2%) |
|  | 2 | 66 (1.8%) | 795 (21.6%) | 977 (14.4%) | 323 (11.5%) | 412 (7%) |
|  | 3 | 184 (5%) | 379 (10.3%) | 1043 (15.3%) | 466 (16.6%) | 461 (7.8%) |
|  | 4 | 303 (8.3%) | 295 (8%) | 846 (12.4%) | 352 (12.5%) | 897 (15.2%) |
|  | 5 | 312 (8.5%) | 174 (4.7%) | 770 (11.3%) | 395 (14.1%) | 658 (11.2%) |
|  | 6 | 422 (11.5%) | 148 (4%) | 556 (8.2%) | 433 (15.4%) | 678 (11.5%) |
|  | 7 | 486 (13.2%) | 169 (4.6%) | 364 (5.4%) | 199 (7.1%) | 793 (13.5%) |
|  | 8 | 687 (18.7%) | 183 (5%) | 247 (3.6%) | 139 (5%) | 531 (9%) |
|  | 9 | 679 (18.5%) | 192 (5.2%) | 140 (2.1%) | 92 (3.3%) | 341 (5.8%) |
|  | 10 | 405 (11%) | 186 (5.1%) | 121 (1.8%) | 98 (3.5%) | 415 (7%) |
|  | NA | 114 (3.1%) | 114 (3.1%) | 207 (3%) | 59 (2.1%) | 223 (3.8%) |
| Baseline comorbidities, No. % | Myocardial infarction | 200 (5.5%) | 152 (4.1%) | 229 (3.4%) | 108 (3.8%) | 422 (7.2%) |
|  | Congestive heart failure | 343 (9.3%) | 324 (8.8%) | 376 (5.5%) | 188 (6.7%) | 760 (12.9%) |
|  | Cerebrovascular disease | 273 (7.4%) | 212 (5.8%) | 357 (5.2%) | 160 (5.7%) | 422 (7.2%) |
|  | Dementia | 26 (0.7%) | 36 (1%) | 42 (0.6%) | 30 (1.1%) | 88 (1.5%) |
|  | Chronic pulmonary disease | 735 (20%) | 547 (14.9%) | 1209 (17.8%) | 376 (13.4%) | 1167 (19.8%) |
|  | Rheumatologic disease | 150 (4.1%) | 125 (3.4%) | 337 (5%) | 119 (4.2%) | 252 (4.3%) |
|  | Peptic ulcer disease | 154 (4.2%) | 102 (2.8%) | 268 (3.9%) | 87 (3.1%) | 303 (5.1%) |
|  | Mild liver disease | 501 (13.7%) | 566 (15.4%) | 787 (11.6%) | 306 (10.9%) | 1011 (17.2%) |
|  | Hemoplegia or paraplegia | 54 (1.5%) | 28 (0.8%) | 35 (0.5%) | 36 (1.3%) | 78 (1.3%) |
|  | Renal disease | 669 (18.2%) | 840 (22.8%) | 950 (14%) | 483 (17.2%) | 1302 (22.1%) |
|  | AIDS/HIV | 27 (0.7%) | 104 (2.8%) | 67 (1%) | 30 (1.1%) | 304 (5.2%) |
|  | Metastatic solid tumor | Less than 10 | Less than 10 | Less than 10 | Less than 10 | Less than 10 |
|  | Moderate to severe liver disease | 40 (1.1%) | 126 (3.4%) | 97 (1.4%) | 25 (0.9%) | 171 (2.9%) |
|  | Any malignancy | 546 (14.9%) | 768 (20.9%) | 1134 (16.7%) | 478 (17%) | 1117 (19%) |
| Sodium-glucose cotransporter-2 inhibitors |  |  |  |  |  |  |
| New users, No. |  | 3349 | 3293 | 5505 | 3057 | 4038 |
| Sex, No. | Female | 1357 (40.52%) | 1249 (37.93%) | 2096 (38.07%) | 1210 (39.58%) | 1562 (38.68%) |
|  | Male | 1992 (59.48%) | 2044 (62.07%) | 3409 (61.93%) | 1847 (60.42%) | 2476 (61.32%) |
| Age, mean (SD) |  | 64.6 (13.7) | 66.4 (14.3) | 64.7 (14.2) | 63.8 (14.2) | 66.1 (13) |
| Treatment Discontinuation, NO. | Yes | 774 | 751 | 1484 | 821 | 1164 |
|  | No | 2575 | 2542 | 4021 | 2236 | 2874 |
| Baseline healthcare visits, mean (SD) |  | 109.9 (161.9) | 80.9 (121.3) | 95 (154.5) | 62.8 (127.3) | 100.4 (164.8) |
| Area deprivation index, No. % | 1 | 11 (0.3%) | 914 (27.8%) | 1273 (23.1%) | 193 (6.3%) | 378 (9.4%) |
|  | 2 | 51 (1.5%) | 809 (24.6%) | 743 (13.5%) | 245 (8%) | 299 (7.4%) |
|  | 3 | 154 (4.6%) | 309 (9.4%) | 819 (14.9%) | 479 (15.7%) | 319 (7.9%) |
|  | 4 | 209 (6.2%) | 255 (7.7%) | 643 (11.7%) | 396 (13%) | 611 (15.1%) |
|  | 5 | 237 (7.1%) | 135 (4.1%) | 559 (10.2%) | 509 (16.7%) | 408 (10.1%) |
|  | 6 | 331 (9.9%) | 130 (3.9%) | 468 (8.5%) | 512 (16.7%) | 484 (12%) |
|  | 7 | 385 (11.5%) | 152 (4.6%) | 300 (5.4%) | 323 (10.6%) | 501 (12.4%) |
|  | 8 | 576 (17.2%) | 179 (5.4%) | 225 (4.1%) | 164 (5.4%) | 338 (8.4%) |
|  | 9 | 798 (23.8%) | 181 (5.5%) | 141 (2.6%) | 70 (2.3%) | 240 (5.9%) |
|  | 10 | 496 (14.8%) | 146 (4.4%) | 129 (2.3%) | 110 (3.6%) | 300 (7.4%) |
|  | NA | 101 (3%) | 83 (2.5%) | 205 (3.7%) | 56 (1.8%) | 160 (4%) |
| Baseline comorbidities, No. % | Myocardial infarction | 785 (23.4%) | 550 (16.7%) | 704 (12.8%) | 415 (13.6%) | 634 (15.7%) |
|  | Congestive heart failure | 1469 (43.9%) | 1256 (38.1%) | 1479 (26.9%) | 803 (26.3%) | 1465 (36.3%) |
|  | Cerebrovascular disease | 475 (14.2%) | 441 (13.4%) | 546 (9.9%) | 316 (10.3%) | 500 (12.4%) |
|  | Dementia | 71 (2.1%) | 98 (3%) | 104 (1.9%) | 80 (2.6%) | 98 (2.4%) |
|  | Chronic pulmonary disease | 1044 (31.2%) | 629 (19.1%) | 894 (16.2%) | 421 (13.8%) | 925 (22.9%) |
|  | Rheumatologic disease | 154 (4.6%) | 126 (3.8%) | 215 (3.9%) | 85 (2.8%) | 168 (4.2%) |
|  | Peptic ulcer disease | 198 (5.9%) | 131 (4%) | 221 (4%) | 108 (3.5%) | 230 (5.7%) |
|  | Mild liver disease | 393 (11.7%) | 498 (15.1%) | 523 (9.5%) | 314 (10.3%) | 498 (12.3%) |
|  | Hemoplegia or paraplegia | 79 (2.4%) | 64 (1.9%) | 57 (1%) | 79 (2.6%) | 67 (1.7%) |
|  | Renal disease | 1402 (41.9%) | 1555 (47.2%) | 1786 (32.4%) | 909 (29.7%) | 1599 (39.6%) |
|  | AIDS/HIV | 31 (0.9%) | 59 (1.8%) | 42 (0.8%) | 26 (0.9%) | 89 (2.2%) |
|  | Metastatic solid tumor | Less than 10 | Less than 10 | Less than 10 | Less than 10 | Less than 10 |
|  | Moderate to severe liver disease | 116 (3.5%) | 168 (5.1%) | 147 (2.7%) | 87 (2.8%) | 146 (3.6%) |
|  | Any malignancy | 710 (21.2%) | 871 (26.5%) | 1223 (22.2%) | 540 (17.7%) | 1010 (25%) |
| Sulfonylureas |  |  |  |  |  |  |
| New users, No. |  | 6756 | 4383 | 5992 | 6033 | 4848 |
| Sex, No. | Female | 2934 (43.43%) | 1901 (43.37%) | 2569 (42.87%) | 2863 (47.46%) | 2116 (43.65%) |
|  | Male | 3822 (56.57%) | 2482 (56.63%) | 3423 (57.13%) | 3170 (52.54%) | 2732 (56.35%) |
| Age, mean (SD) |  | 61.3 (12.4) | 62 (12.4) | 62.7 (13) | 60.5 (13.5) | 61.4 (12.4) |
| Treatment Discontinuation, NO. | Yes | 2701 | 1769 | 2504 | 2174 | 2204 |
|  | No | 4055 | 2614 | 3488 | 3859 | 2644 |
| Baseline healthcare visits, mean (SD) |  | 27.3 (49.1) | 28.1 (49.5) | 32.9 (58.9) | 21 (40.9) | 27.3 (57.8) |
| Area deprivation index, No. % | 1 | 43 (0.6%) | 861 (19.6%) | 1097 (18.3%) | 158 (2.6%) | 183 (3.8%) |
|  | 2 | 143 (2.1%) | 1050 (24%) | 684 (11.4%) | 257 (4.3%) | 221 (4.6%) |
|  | 3 | 244 (3.6%) | 443 (10.1%) | 795 (13.3%) | 663 (11%) | 269 (5.5%) |
|  | 4 | 415 (6.1%) | 401 (9.1%) | 681 (11.4%) | 787 (13%) | 702 (14.5%) |
|  | 5 | 485 (7.2%) | 215 (4.9%) | 716 (11.9%) | 1042 (17.3%) | 560 (11.6%) |
|  | 6 | 606 (9%) | 221 (5%) | 618 (10.3%) | 1409 (23.4%) | 599 (12.4%) |
|  | 7 | 868 (12.8%) | 238 (5.4%) | 417 (7%) | 725 (12%) | 772 (15.9%) |
|  | 8 | 1134 (16.8%) | 270 (6.2%) | 344 (5.7%) | 425 (7%) | 599 (12.4%) |
|  | 9 | 1567 (23.2%) | 301 (6.9%) | 214 (3.6%) | 183 (3%) | 269 (5.5%) |
|  | 10 | 981 (14.5%) | 245 (5.6%) | 179 (3%) | 266 (4.4%) | 413 (8.5%) |
|  | NA | 270 (4%) | 138 (3.1%) | 247 (4.1%) | 118 (2%) | 261 (5.4%) |
| Baseline comorbidities, No. % | Myocardial infarction | 361 (5.3%) | 226 (5.2%) | 309 (5.2%) | 337 (5.6%) | 286 (5.9%) |
|  | Congestive heart failure | 626 (9.3%) | 459 (10.5%) | 451 (7.5%) | 497 (8.2%) | 548 (11.3%) |
|  | Cerebrovascular disease | 471 (7%) | 253 (5.8%) | 289 (4.8%) | 392 (6.5%) | 246 (5.1%) |
|  | Dementia | 136 (2%) | 98 (2.2%) | 130 (2.2%) | 162 (2.7%) | 113 (2.3%) |
|  | Chronic pulmonary disease | 745 (11%) | 389 (8.9%) | 526 (8.8%) | 379 (6.3%) | 475 (9.8%) |
|  | Rheumatologic disease | 130 (1.9%) | 85 (1.9%) | 124 (2.1%) | 96 (1.6%) | 103 (2.1%) |
|  | Peptic ulcer disease | 130 (1.9%) | 86 (2%) | 124 (2.1%) | 95 (1.6%) | 135 (2.8%) |
|  | Mild liver disease | 436 (6.5%) | 599 (13.7%) | 468 (7.8%) | 337 (5.6%) | 440 (9.1%) |
|  | Hemoplegia or paralegia | 103 (1.5%) | 83 (1.9%) | 56 (0.9%) | 126 (2.1%) | 63 (1.3%) |
|  | Renal disease | 1502 (22.2%) | 1401 (32%) | 1446 (24.1%) | 1070 (17.7%) | 1007 (20.8%) |
|  | AIDS/HIV | 21 (0.3%) | 68 (1.6%) | 33 (0.6%) | 44 (0.7%) | 131 (2.7%) |
|  | Metastatic solid tumor | Less than 10 | Less than 10 | Less than 10 | Less than 10 | Less than 10 |
|  | Moderate to severe liver disease | 64 (0.9%) | 281 (6.4%) | 116 (1.9%) | 87 (1.4%) | 169 (3.5%) |
|  | Any malignancy | 767 (11.4%) | 917 (20.9%) | 907 (15.1%) | 689 (11.4%) | 727 (15%) |

Treatment discontinuation rates also differed. DPP-4i and sulfonylurea had the highest discontinuation rates (39%–46% and 36%–45%, respectively), suggesting frequent use as initial therapies before transitioning. In contrast, GLP-1RA users demonstrated lower discontinuation rates (16%–21%). Renal disease was the most prevalent comorbidity, with the highest burden among SGLT-2i users (30%–47%), followed by sulfonylurea users (18%–32%), and the lowest among GLP-1RA users (14%–23%). Heart failure was most common in SGLT-2i users (26%–44%). Malignancy prevalence ranged from 11%–26% across all classes, reflecting the older age of the cohort.

### Matched population and phenome-wide association

A total of 15 Tc–Cc cohort pairs were analyzed, encompassing pairwise combinations among the four drug classes (GLP-1RA, sulfonylurea, SGLT-2i, and DPP-4i) across five UC Health sites. Propensity scores were computed using a high-dimensional feature set that included a mean (SD) of 6,093 (513.23) pretreatment covariates, in addition to patient age at treatment initiation, sex, area deprivation index (ADI) as a proxy for socioeconomic status, and number of visits as healthcare utilization indicators at each site.

For example, **Figure 2** displays the propensity score distributions for patients treated with GLP-1RA versus sulfonylurea at each site independently. For instance, at UC-2 **(Figure-2 B)** propensity scores were estimated using 6,211 covariates, including 1,017 procedure codes, 1,528 laboratory measurements, 2,107 medication orders, and 1,559 diagnostic codes, along with demographic and socioeconomic covariates captured on or within 365 days prior to the index date. Post-matching, 99% of covariates achieved balance, defined as an absolute standardized mean difference (SMD) < 0.10, resulting in a matched analytic cohort of 2,848 patients **(Figure-2 B)**. The number of pre-treatment covariates used in propensity score estimation and number of covariates balanced before and after matching for all 15 Tc–Cc comparisons across all five sites are provided in **eTable 2.**

**Figure 2:**
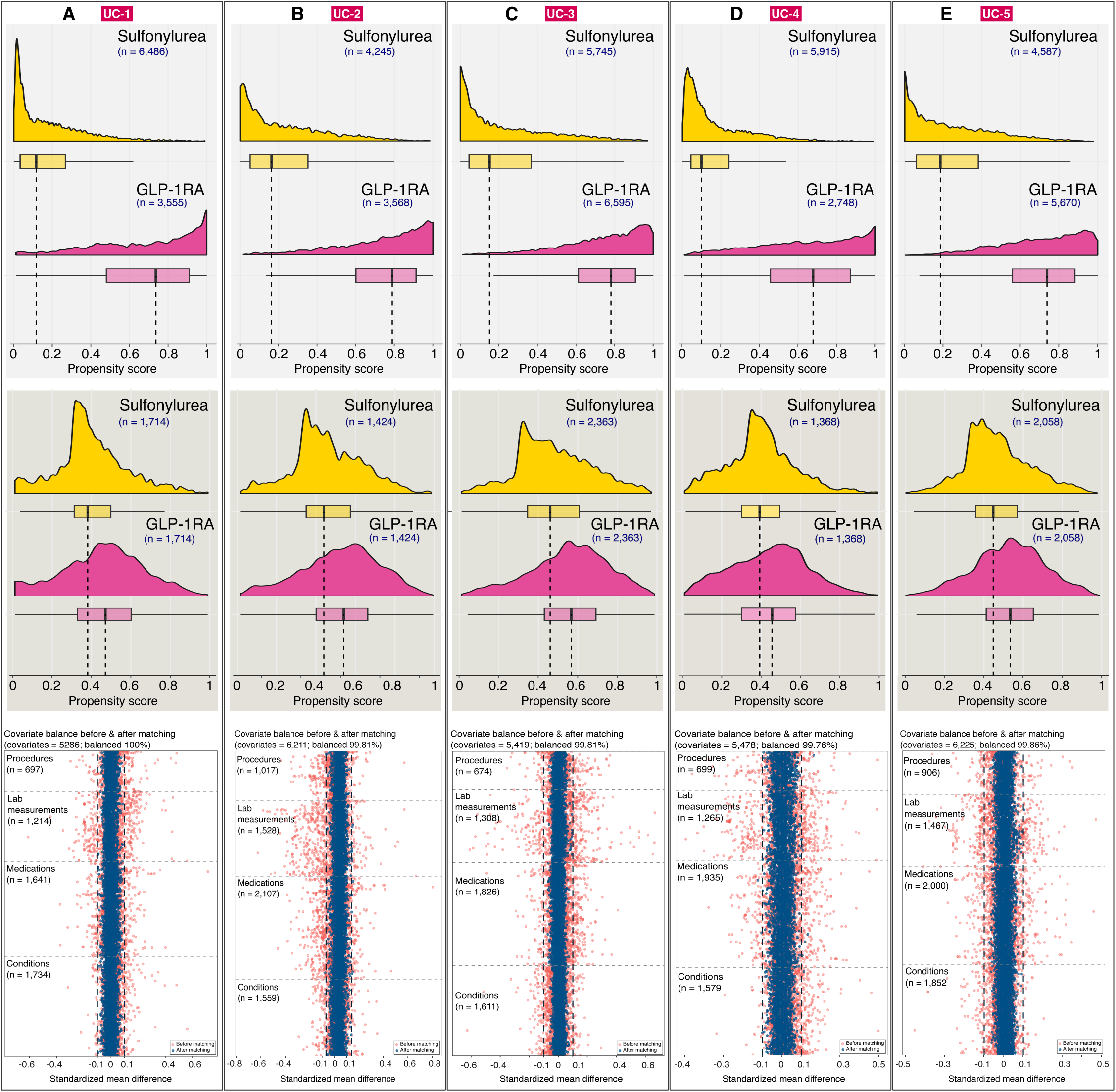
Propensity score distribution and balance diagnostics. Distribution of propensity scores before and after matching as well as covariate balance diagnostic across UC Health for sulfonylurea vs GLP1-RA comparison. A) (top panel) propensity score distribution before matching, (middle panel) propensity score distribution after matching, and (bottom panel) covariate balance in terms of standardized mean difference of 5,286 covariates used in the propensity score estimation at UC-1.

A total of 239 outcomes from 14 distinct body systems were included, mean 17 phenotypes per system **(Figure 3)**, high 34 (circulatory system) to low 6 (ear and mastoid process). In the sulfonylurea group across UC Health, 235 of 239 phenotypes qualified for analysis; 167 (71.1%) from all five UCs. In the DPP4i group, 230 of 239 phenotypes qualified; estimates for 156 (67.8%) were derived from all five UCs. In the SGLT-2i group, 228 of 239 phenotypes qualified; 158 (69.3%) phenotypes had estimates derived from all five UCs. Across all cohorts, most phenotypes (67.8% to 71.1%) had risk estimates reproducibly derived from all five UCs, demonstrating consistent data capture and phenotype qualification **(Figure 3);** this reproducibility pertains solely to the estimation process and does not indicate uniformity in the magnitude, direction, or statistical significance of the risk effects. The estimated effect size, along with 95% CI and p-values along with number of events of each clinical outcome by the body system across all the UCs and for each comparison is provided in **eTable 3**.

**Figure 3:**
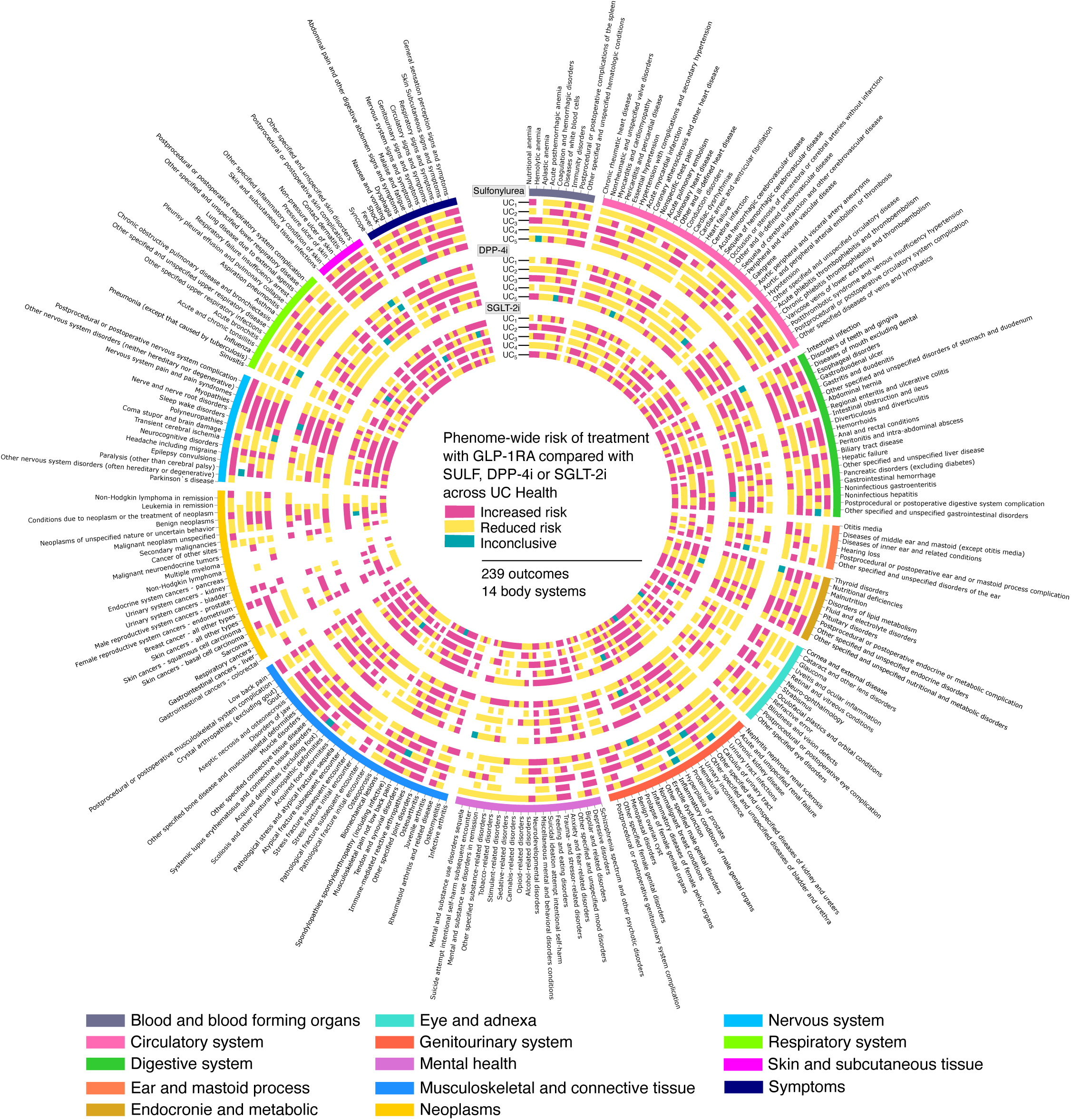
Phenome-wide associations. Elevated or reduced risk of 239 clinical phenotypes on treatment GLP-1RA compared with sulfonylurea, DPP-4i or SGL-2i across 5 UCs. The outermost ring delineates phenotypic categories (clinical endpoints, n = 239) stratified by body system (n = 14). Concentric rings represent hazard ratios for increased or reduced risk of corresponding clinical endpoints within each organ system for sulfonylurea, DPP-4i and SGLT-2i compared with treatment GLP-1RA at each of the UC.

### Real-World Evidence Landscape of Effect of GLP-1RA Treatment

A total of 32 clinical endpoints demonstrated a significantly reduced risk, while 25 clinical endpoints exhibited a significantly increased risk associated with GLP-1RA treatment compared to either sulfonylurea, DPP- 4i, or SGLT-2i, evaluated independently **(Figure 4 A, B)**. All 32 phenotypes with reduced risk met criteria for stability as defined by LOMCO stability analysis and low heterogeneity (I² < 20%), indicating consistency across UC sites. Among the phenotypes associated with increased risk, 24 clinical endpoints met this stability criteria. After applying corrections for multiple hypothesis testing and assessing susceptibility to unmeasured confounding, 18 phenotypes among those initially linked to reduced risk and 10 phenotypes among those linked to increased risk retained statistical significance (**eTable 4)**.

**Figure 4:**
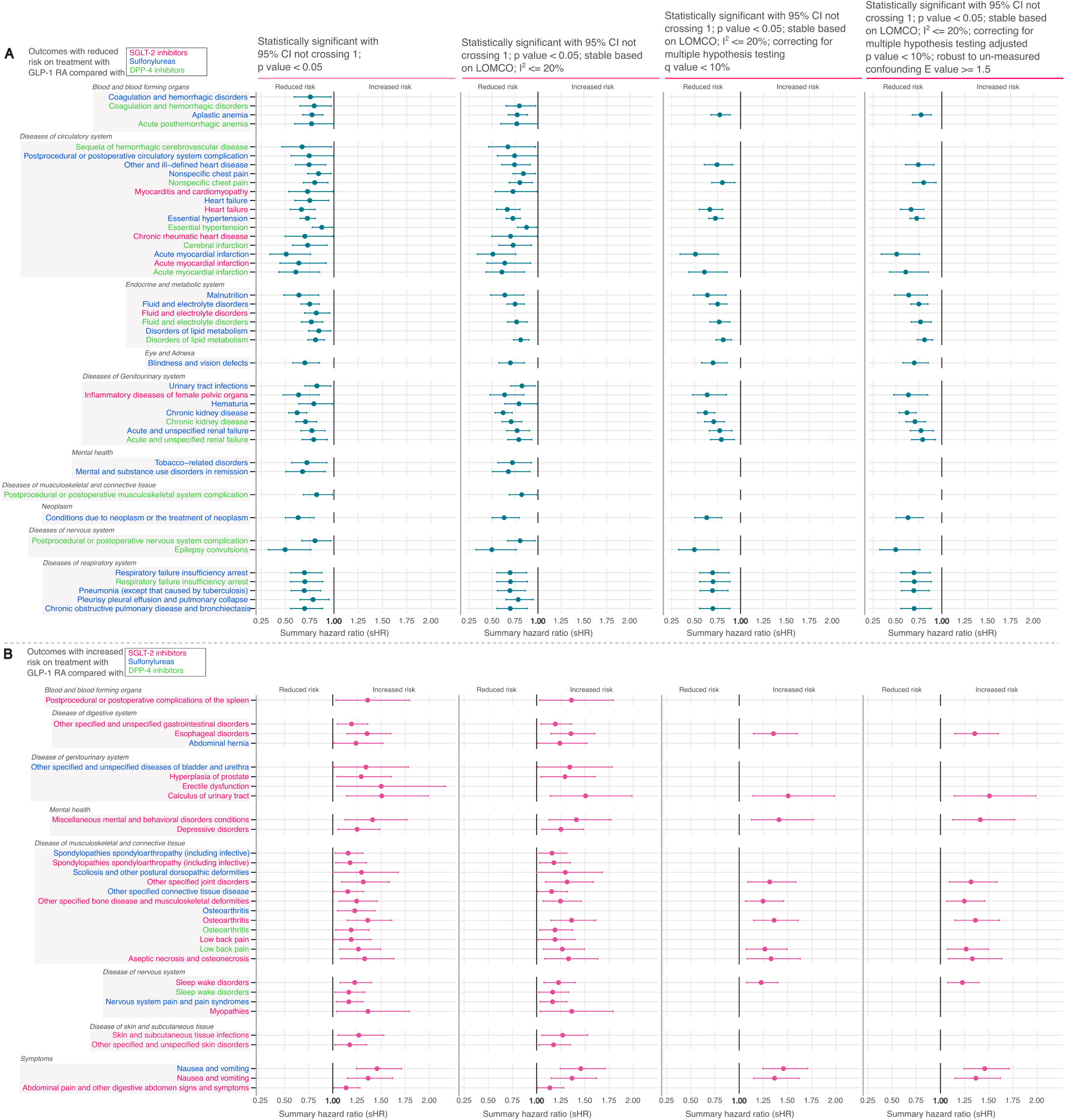
Real-world evidence landscape at varying degree of statistical significance. Evidence landscape of comparative risk profiles at varying degree of statistical certainty, significance and un-measured confounding comparing GLP-1RA (treatment) and sulfonylurea, SGLT-2i, or DPP-4i (controls). A) Outcomes with reduced risk B) outcomes with increased risk.

### Phenotypes with reduced risk on treatment with GLP-1RA Cardiovascular outcomes

Treatment with GLP-1RA was associated with reduced risk of acute myocardial infarction versus DPP4i (sHR 0.61, 95% CI 0.43–0.85) and sulfonylurea (sHR 0.51, 95% CI 0.34–0.76) **(Figure-4 A)**.

Furthermore, GLP1-RA treatment was associated with a reduced incidence of heart failure compared to SGLT-2i (sHR 0.66, 95% CI 0.55–0.80) and hypertension relative to sulfonylurea (sHR 0.72, 95% CI 0.65–0.80).

### Renal outcomes

Treatment with GLP-1RA was associated with reduced risk of chronic kidney disease versus DPP- 4i (sHR 0.71, 95% CI 0.60-0.82) and sulfonylurea (sHR 0.62, 95% CI 0.53-0.72), while acute renal failure risk decreased (sHR 0.79, 95% CI 0.67–0.93) (sHR 0.77, 95% CI 0.65–0.90) compared to DPP-4i and sulfonylurea, respectively **(Figure-4 A)**.

### Respiratory outcomes

Treatment with GLP1 was also associated with lower risk of chronic obstructive pulmonary disease (sHR 0.69, 95% CI 0.55–0.88), pneumonia (sHR 0.69, 95% CI 0.55–0.86), and respiratory failure or insufficiency (sHR 0.69, 95% CI 0.55–0.87) compared to sulfonylurea. GLP-1RA was also associated with a reduced risk of epilepsy or seizures versus DPP-4i (sHR 0.49, 95% CI 0.32–0.76) and of blindness or vision defects compared to sulfonylureas (sHR 0.70, 95% CI 0.57–0.85) **(Figure-4 A)**.

### Phenotypes with increased risk on treatment with GLP-1RA Musculoskeletal and Connective Tissue

Treatment with GLP-1RA was associated with elevated risk of musculoskeletal outcomes compared to other antidiabetic therapies, particularly SGLT-2i **(Figure-4 B)**. Musculoskeletal conditions with higher hazards among GLP-1RA users included low back pain (HR 1.27, 95% CI: 1.07–1.50), osteoarthritis (HR 1.36, 95% CI: 1.15–1.61), bone diseases and musculoskeletal deformities (sHR 1.24, 95% CI: 1.12–1.76), and aseptic necrosis (sHR 1.33, 95% CI: 1.08–1.63) relative to SGLT-2i users.

### Digestive System

The hazard for esophageal disorders was elevated with GLP-1RA compared to SGLT-2i (sHR 1.36, 95% CI: 1.15–1.60). Gastrointestinal symptoms, particularly nausea and vomiting were observed more frequently relative to both SGLT-2i (sHR 1.37, 95% CI 1.15–1.62) and sulfonylurea (sHR 1.46, 95% CI 1.24–1.71) **(Figure-4 B)**.

### Nervous System and Genitourinary system

Within the nervous system domain, GLP-1RA use was associated with a higher hazard of sleep-wake disorders compared to SGLT-2i (sHR 1.23, 95% CI 1.07-1.40). Additionally, in the genitourinary domain, GLP1RA was associated with an increased hazard of urinary tract calculus (sHR 1.51, 95% CI 1.14-1.98 vs. SGLT-2i) **(Figure-4 B)**.

## Discussion

Our comparison of GLP-1RA with other therapies among a large, real-world cohort of patients with T2D verified efficacy and adverse effect profiles that have been well-established by RCTs, supporting the credibility of using routine clinical data for robust, phenome-wide investigations New users of SGLT-2 inhibitors had a greater burden of renal disease and congestive heart failure compared with new users of other glucose-lowering therapies—findings that align with guideline-driven preferential use of SGLT-2i in these high-risk populations^30^. Treatment switching patterns further illuminate prescriber and patient preferences. The high switching rates from sulfonylureas (36%–45%) and DPP-4i (39%– 46%) suggest intolerance, consistent with their lower efficacy and higher hypoglycemia risk compared to newer agents^31–33^. However, switching may also reflect clinician-driven medication changes in response to emerging evidence and evolving treatment guidelines. Conversely, the durability of GLP-1 RA therapy (switching rates 16%–21%) aligns with studies demonstrating superior patient satisfaction and adherence, particularly when weight loss or cardiovascular benefits are perceived^34,35^. However, recent studies have identified higher rates of GLP-1RA discontinuation, underscoring the dynamic and often cyclical nature of GLP-1 RA utilization in real-world clinical practice^36^.

Given the intriguing and sometimes heterogeneous patterns observed between treatment choice and comorbid conditions, we extended our analysis to a broader phenome-wide association study. Our goal was to capture a more comprehensive view of the real-world impact of glucose-lowering therapies across diverse clinical contexts. By systematically comparing GLP-1RA with other commonly used therapies (sulfonylureas, SGLT-2i, and DPP-4i), we aimed to test the robustness of known associations, uncover novel signals – including safety signals that require larger sample sizes than in clinical trials, and evaluate the utility of routinely collected clinical data for supporting broad pharmacophenomic investigations.

Consistent with RCT evidence, we observed cardiorenal benefits of GLP-1 RA. GLP-1 RAs reduce atherosclerotic plaque burden through anti-inflammatory and endothelial-stabilizing effects^37^, which may explain the 39.3% and 49.2% risk reductions in acute myocardial infarction compared to DPP-4i and sulfonylureas, respectively. These results extend findings from cardiovascular outcome trials including LEADER^5^ and SUSTAIN- 6^6^, which demonstrated MACE reduction in high-risk populations^38^, to real-world cohorts with broader cardiovascular risk profiles. Notably, the 34% lower heart failure risk versus sodium-glucose cotransporter-2 inhibitors (SGLT2i) challenges conventional hierarchies in diabetes care, suggesting complementary mechanisms—such as GLP-1 RA–mediated improvements in ventricular filling pressures^37^—that warrant further investigation. Renoprotective effects, including a 29–38% risk reduction in chronic kidney disease compared to DPP-4i and sulfonylureas, mirror the hemodynamic benefits observed in trials such as REWIND^35^ and FLOW^6^, where GLP-1 RA attenuated albuminuria progression among other clinical benefits. The consistency of these results across heterogeneous real-world populations not only strengthens the case for early GLP-1 RA initiation in patients with incipient renal dysfunction but also underscores the credibility of real-world data for informing therapeutic decisions.

The metabolic advantages of GLP-1 RAs extend beyond glycemic control. Respiratory benefits, including lower risks of chronic obstructive pulmonary disease and pneumonia, could be mediated by anti-inflammatory actions. Potential neurological benefits associated with GLP-1 receptor agonist treatment include lower rates of seizure-related events, as well as reductions in visual impairments, based on direct head-to-head comparisons with other therapies. These findings build upon and strengthen emerging evidence from recent meta-analyses of RCTs, which have suggested similar effects but were oen limited by the absence of such direct comparisons^39^. Collectively, these results support novel hypotheses regarding neuroprotective mechanisms potentially mediated via GLP-1 receptor signaling in the central nervous system. GLP-1RAs are being investigated for potential benefits in neurodegenerative conditions such as dementia, and seizure-related events may, in some cases, be secondary to cerebrovascular insults such as stroke—an area where GLP-1RAs have demonstrated cardiovascular benefit^40^.

These findings certainly warrant further investigation in dedicated studies before drawing definitive conclusions about the neurological implications of GLP-1RA therapy.

While GLP-1RAs demonstrated robust efficacy, our analysis identified underappreciated risks. Musculoskeletal outcomes, including a 26–36% elevated risk of low back pain and osteoarthritis versus SGLT2i, may relate to rapid weight loss–induced biomechanical changes or direct effects on bone metabolism^41,42^. While randomized controlled trial evidence demonstrates that GLP-1 receptor agonists can reduce osteoarthritis-related pain^14^, our findings suggest that new diagnoses of osteoarthritis may nonetheless be more common following GLP-1RA use. This observation aligns with prior reports that raise similar concerns^19,43^. One potential explanation is that patients may experience increased mechanical loading on joints due to greater physical activity following weight loss. Thus, while acknowledging the therapeutic benefits of GLP-1RAs for osteoarthritis symptom management, our data highlight the need to carefully consider possible biomechanical consequences and the complex relationship between weight loss, joint health, and physical activity.

Our study was able to capture the well-established gastrointestinal adverse events^3,44^. The underreported association with genitourinary conditions, such as urinary tract calculi, raises questions about dehydration risks or altered mineral excretion, necessitating mechanistic studies. Mental health risks, including sleep-wake disorders, contrast with preclinical data suggesting GLP-1 RA neuroprotection. This discrepancy may reflect real-world confounding (e.g., psychological stressors in obese populations) or off-target effects of prolonged receptor agonism, highlighting the need for dedicated pharmacovigilance studies. These findings complement and build upon evidence emerging from large-scale pharmacoepidemiologic resources such as the atlas of health associations^19^, while also extending their insights by uncovering novel and underappreciated safety signals across diverse populations and healthcare systems. By leveraging real-world data and rigorous comparative analyses, our study underscores the indispensable role of post-marketing surveillance and mechanistic follow-up studies in refining the safety profile of GLP-1 RAs in everyday clinical practice as these medications are more widely used.

As GLP-1 receptor agonists (GLP-1RAs) are adopted for an expanding range of indications and patient populations, robust post-market surveillance leveraging real-world data from electronic health records will be essential. Target trial emulation frameworks, as implemented in this study, facilitate timely hypothesis generation and iterative evaluation of safety and effectiveness, aligning with the increasing role and potential of such observational data analytic approaches in both clinical practice and regulatory decision-making^45^. Ultimately, the integration of high-fidelity observational evidence with RCT data will support precise, patient-centered monitoring and risk–benefit assessment, ensuring the safe and effective deployment of GLP-1RAs across diverse clinical settings.

## Limitations

Our study had limitations. Our study population includes patients receiving care at any of the UC Health facilities across California. Patients within the UC Health system may receive care outside of UC Health, such as hospitalization for an acute event. However, the likelihood of such occurrences is low, as most patients in our study population receive comprehensive care within UC Health. Moreover, we do not anticipate that the probability of seeking outside care would differ systematically by treatment group; thus, any resulting outcome misclassification would likely be non-differential and unlikely to bias the comparative effectiveness estimates. Nonetheless, the broader issue of fragmented care—where patients receive services across multiple, unconnected health systems—presents both a limitation of our study and a challenge in generalizing findings to health systems beyond California. We did not include race, ethnicity, or treatment cost in our analysis, primarily due to limitations in the availability and completeness of these data within EHRs. In particular, race and ethnicity are often missing or inconsistently recorded, especially in outpatient and ambulatory settings. These gaps, along with challenges in capturing patients’ financial contexts, limited our ability to account for these important factors in the current study.

Obesity was not included as an outcome phenotype in this study due to its potential role influencing treatment assignment. We observed a substantial number of new obesity diagnoses occurring within 7 to 14 days following the index date, which likely reflect post-hoc coding decisions made after the initiation of therapy— particularly given the regulatory approval of GLP-1 receptor agonists for obesity management. As such, these diagnoses may not represent incident cases of obesity following treatment initiation but rather retrospective documentation, thereby limiting their validity as treatment-emergent outcomes within this narrow temporal window. However, obesity diagnoses present prior to treatment assignment were adjusted for in the propensity score estimation.

## Conclusion

This multi-center study synthesizes real-world evidence from diverse healthcare systems to evaluate the broader safety profile of GLP-1RAs beyond the primary outcomes that supported their initial approval. With the rapid rise in GLP-1RA utilization across a range of indications, it is imperative to rigorously assess their long-term risk-benefit profile. Our findings both align with existing evidence from RCTs while also contributing new knowledge to the growing body of real-world data that underscores the importance of continued safety monitoring. As GLP-1RAs increasingly emerge as multifaceted agents beyond glycemic control, sustained real-world evidence will be essential for optimizing their therapeutic use across varied populations. Such evidence can inform hypothesis generation, refine the design of future RCTs, and support more precise, actionable clinical decision-making.

## Supporting information

eTable-1

eTable-2

eTable-3

eTable-4

## Data Availability

The data used for this analysis was derived from the de-identified electronic health records of patients receiving care across the University of California Health. Although de-identified, the individual-level nature of the data used risks individuals being identified, or being able to self-identify, if the data are released publicly.

## Funding

This research received primary support from the Clinical and Translational Science Institute at the University of California, San Francisco (UCSF), under award number UL1TR001872. Additional funding was partially provided by the U.S. Food and Drug Administration (FDA) through grant number U01FD005978, awarded to the UCSF–Stanford Center of Excellence in Regulatory Science and Innovation. Further support was contributed by the UCSF Bakar Computational Health Sciences Institute and the University of California Health Center for Data-driven Insights and Innovation.

## Role of Funders

The funding agencies had no involvement in the design or execution of the study; the collection, management, analysis, or interpretation of the data; the preparation, review, or approval of the manuscript; or the decision to submit it for publication.

## Acknowledgments

We thank Ana M. Deluca, Emrica A. Pitolin, and Andenet Emiru at UCSF and University of California Health for their assistance with the UC Health Data Warehouse. There contributions were made as part of their regular employment and were not separately compensated.

## Deceleration of interests

RV, AP, LD, PM, DG, RF, CH, MS have no conflict of interests. KS is a consultant for Google and serves on their consumer health advisory panel. AB reported serving as a cofounder and consultant to Personalis and NuMedii; serving as a consultant to Mango Tree Corporation, Samsung, 10x Genomics, Helix, Pathway Genomics, and Verinata (Illumina); serving on paid advisory panels or boards for Geisinger Health, Regenstrief Institute, Gerson Lehman Group, AlphaSights, Covance, Novartis, Genentech, and Merck, and Roche; owning stock in Personalis, NuMedii, Apple, Meta, Alphabet, Microsoft, Amazon, Snap, 10x Genomics, Illumina, Regeneron, Sanofi, Pfizer, Royalty Pharma, Moderna, Sutro, Doximity, BioNtech, Invitae, Pacific Biosciences, Editas Medicine, Nuna Health, Assay Depot, and Vet24seven; receiving personal fees from Johnson and Johnson, Roche, Genentech, Pfizer, Merck, Lilly, Takeda, Varian, Mars, Siemens, Optum, Abbott, Celgene, AstraZeneca, AbbVie, Westat, Stanford University, NuMedii, Personalis, Dartmouth University, Boston Children’s Hospital, Johns Hopkins University, Endocrine Society, Alliance for Academic Internal Medicine, Children’s Hospital of Philadelphia, University of Pittsburgh Medical Center, Cleveland Clinic, University of Utah, Society of Toxicology, Mayo Clinic, Washington University in Saint Louis, and University of Michigan; receiving grants from the National Institutes of Health, Peraton, Genentech, Johnson and Johnson, US Food and Drug Administration (FDA), Robert Wood Johnson Foundation, Leon Lowenstein Foundation, Intervalien Foundation, Priscilla Chan and Mark Zuckerberg, the Barbara and Gerson Bakar Foundation, March of Dimes, Juvenile Diabetes Research Foundation, California Governor’s Office of Planning and Research, California Institute for Regenerative Medicine, L’Oréal, and Progenity outside the submitted work. No other disclosures were reported.

## Notes

### Author Declarations

The Institutional Review Boards across the University of California Health system determined that the research use of the HIPAA limited data set for this cohort study did not constitute human participants research and was therefore exempt from further approval and informed consent.

